# First Clinical Use of Lenzilumab to Neutralize GM-CSF in Patients with Severe and Critical COVID-19 Pneumonia

**DOI:** 10.1101/2020.06.08.20125369

**Authors:** Zelalem Temesgen, Mariam Assi, Paschalis Vergidis, Stacey A. Rizza, Philippe R. Bauer, Brian W. Pickering, Raymund R. Razonable, Claudia R. Libertin, Charles D. Burger, Robert Orenstein, Hugo E. Vargas, Bharath Raj Varatharaj Palraj, Ala S. Dababneh, Gabrielle Chappell, Dale Chappell, Omar Ahmed, Reona Sakemura, Cameron Durrant, Saad S. Kenderian, Andrew D. Badley

## Abstract

**Background:** In COVID-19, high levels of granulocyte macrophage-colony stimulating factor (GM-CSF) and inflammatory myeloid cells correlate with disease severity, cytokine storm, and respiratory failure. With this rationale, we used lenzilumab, an anti-human GM-CSF monoclonal antibody, to treat patients with severe and critical COVID-19 pneumonia.

**Methods:** Hospitalized patients with COVID-19 pneumonia and risk factors for poor outcomes were treated with lenzilumab 600 mg intravenously for three doses through an emergency single-use IND application. Patient characteristics, clinical and laboratory outcomes, and adverse events were recorded. All patients receiving lenzilumab through May 1, 2020 were included in this report.

**Results:** Twelve patients were treated with lenzilumab. Clinical improvement was observed in 11 out of 12 (92%), with a median time to discharge of 5 days. There was a significant improvement in oxygenation: The proportion of patients with SpO2/FiO2 < 315 at the end of observation was 8% vs. compared to 67% at baseline (p=0.00015). A significant improvement in mean CRP and IL-6 values on day 3 following lenzilumab administration was also observed (137.3 mg/L vs 51.2 mg/L, p = 0.040; 26.8 pg/mL vs 16.1 pg/mL, p = 0.035; respectively). Cytokine analysis showed a reduction in inflammatory myeloid cells two days after lenzilumab treatment. There were no treatment-emergent adverse events attributable to lenzilumab, and no mortality in this cohort of patients with severe and critical COVID-19 pneumonia.

**Conclusions:** In high-risk COVID-19 patients with severe and critical pneumonia, GM-CSF neutralization with lenzilumab was safe and associated with improved clinical outcomes, oxygen requirement, and cytokine storm.

## INTRODUCTION

The clinical manifestations of COVID-19, the disease caused by severe acute respiratory coronavirus 2 (SARS-CoV-2) infection, range from asymptomatic disease to severe and critical pneumonia. ^1,2^ Although viral evasion of host immune response and virus-induced cytopathic effects are believed to be critical for disease progression, most deaths associated with COVID-19 are attributed to the development of cytokine release syndrome (CRS) and resultant acute respiratory distress syndrome (ARDS). ^3^

CRS is characterized by an elevation of inflammatory cytokines resulting in fever, hypotension, capillary leak syndrome, pulmonary edema, disseminated intravascular coagulation, respiratory failure, and ARDS. ^4,5^ The development of CRS as a direct result of immune hyper-stimulation has been previously described in patients with autoimmune and lymphoproliferative diseases,^6^ as well as in patients with B-cell malignancies receiving chimeric antigen receptor T-cell (CART) therapy. ^7,8^ Over the last five years, preclinical studies and correlative science from clinical trials in CART therapy have shed light on the pathophysiology, development, characterization, and management of CRS. ^5,9^

CRS during CART therapy is characterized by activation of myeloid cells and release of inflammatory cytokines, including interleukin-6 (IL-6), granulocyte-monocyte colony stimulating factor (GM-CSF), monocyte chemoattractant protein −1 (MCP-1), macrophage inflammatory protein 1α (MIP-1α), Interferon gamma-induced protein 10 (IP-10), and interleukin-1 (IL-1). ^4,7,10^The cascade, once initiated, can quickly evolve into a cytokine storm, resulting in further activation, expansion and trafficking of myeloid cells, leading to abnormal endothelial activation, increased vascular permeability, and disseminated intravascular coagulation. ^11,12^

Similar to patients receiving CART therapy, the development of CRS in patients with COVID-19 has been associated with elevation of CRP, ferritin, and IL-6, as well as correlating with respiratory failure, ARDS, and adverse clinical outcomes.^13-17^ Most significantly, high levels of GM-CSF-secreting Th17 T-cells have been associated with disease severity, myeloid cell trafficking to the lungs, and ICU admission.^18^ This indicates that post-COVID-19 CRS is caused by a similar mechanism, induced by activation of myeloid cells and their trafficking to the lung, resulting in lung injury and ARDS.^18^ Tissue CD14+ myeloid cells produce GM-CSF and IL-6, further triggering a cytokine storm cascade.^18^ Single-cell RNA sequencing of bronchoalveolar lavage samples from COVID-19 patients with severe ARDS demonstrated an overwhelming infiltration of newly-arrived inflammatory myeloid cells compared to mild COVID-19 disease and healthy controls, consistent with a hyperinflammatory CRS-mediated pathology.^19^

With this understanding of the pathophysiology of COVID-19, modalities to target inflammatory cytokines and suppress or prevent CRS after COVID-19 have been investigated in pilot clinical trials. IL-6 blockade has shown encouraging results.^20^ Controlled clinical trials using IL-6 blockade, as well as other immunomodulatory molecules targeting receptor tyrosine kinase are ongoing.

GM-CSF depletion has been developed as a strategy to mitigate CRS following CART therapy. We have shown that GM-CSF neutralization results in a reduction in IL-6, MCP-1, MIP-1α, IP-10, vascular endothelial growth factor (VEGF), and tumor necrosis factor-α (TNFα) levels, demonstrating that GM-CSF is an upstream regulator of many inflammatory cytokines that are important in the pathophysiology of CRS. ^21^ GM-CSF depletion results in modulation of myeloid cell behavior, a specific decrease in their inflammatory cytokines, and a reduction in tissue trafficking, ^21^ while enhancing T-cell apoptosis machinery.^22^ These biological effects prevented both CRS and neuro-inflammation after CART therapy in preclinical models and are being tested in a phase Ib/II clinical trial (NCT 04314843).

Lenzilumab is a first-in-class Humaneered^®^ recombinant monoclonal antibody targeting human GM-CSF, with potential immunomodulatory activity, high binding affinity in the picomolar range, 94% homology to human germline, and has low immunogenicity. Following intravenous administration, lenzilumab binds to and neutralizes GM-CSF, preventing GM-CSF binding to its receptor, thereby preventing GM-CSF-mediated signaling to myeloid progenitor cells.^23^ Lenzilumab has been studied across 4 completed clinical trials in healthy volunteers, and persons with asthma, rheumatoid arthritis, and chronic myelomonocytic leukemia.^24,25^ A total of 113 individuals received lenzilumab in these trials; lenzilumab was very well tolerated with a low frequency and severity of adverse events.^24,25^

Given the hypothesized role of GM-CSF in the pathogenesis of COVID-19 related CRS, along with our studies demonstrating that GM-CSF depletion prevents CRS and modulates myeloid cell behavior in preclinical models,^21^ lenzilumab therapy was offered to patients hospitalized with severe COVID-19 pneumonia, who had clinical and/or biomarker evidence for increased risk of progression to respiratory failure.

## METHODS

### Patients

Hospitalized patients with COVID-19, confirmed by reverse transcriptase-polymerase chain reaction for the SARS-CoV-2, and radiographic findings consistent with COVID-19 pneumonia were considered for treatment with lenzilumab through an emergency IND program. Active systemic infection with bacteria, fungi, or other viruses, was an exclusion criterion. All patients received lenzilumab 600 mg administered via a 1-hour intravenous infusion every 8 hours for a total of three doses (1800 mg). A request for lenzilumab under FDA emergency use IND was submitted to the FDA in accordance with agency guidelines (https://www.fda.gov/regulatory-information/search-fda-guidance-documents/emergency-use-investigational-drug-or-biologic). Informed consent and Institutional review board approval was obtained for each patient.

### Study assessments

There were no pre-specified study endpoints or mandated procedures. All laboratory tests and radiologic assessments were performed at the discretion of the treating physician and per standard clinical management processes. Vital signs were monitored before and upon completion of each lenzilumab infusion. Demographics, co-existing conditions, laboratory and radiographic data, as well as clinical data, adverse events, and outcomes were captured from the electronic health record until data cutoff on May 1, 2020. Data were for all patients for a minimum of five days following the administration of lenzilumab. Baseline values were defined as those values obtained prior to lenzilumab administration, either on the day of administration or the day before the administration. Cytokine analysis was performed on available serum isolated from patients, pre and post lenzilumab treatment. Serum was diluted 1:2 with assay buffer before following the manufacturer’s protocol for Milliplex Human Cytokine/Chemokine MAGNETIC BEAD Premixed 38 Plex Kit (Millipore Sigma, Ontario, Canada). Data were collected using a Luminex (Millipore Sigma, Ontario, Canada).

### Statistical Methods

Continuous variables at baseline are represented using the median and interquartile range (IQR). This is demonstrative of the features in the middle 50% of the cohort. We used an 8-point ordinal outcome scale to define clinical status: 1) Death; 2) Hospitalized, on invasive mechanical ventilation or extracorporeal membrane oxygenation (ECMO); 3) Hospitalized, on non-invasive ventilation or high flow oxygen devices; 4) Hospitalized, requiring supplemental oxygen; 5) Hospitalized, not requiring supplemental oxygen - requiring ongoing medical care (COVID-19 related or otherwise); 6) Hospitalized, not requiring supplemental oxygen - no longer requires ongoing medical care; 7) Not hospitalized, limitation on activities; 8) Not hospitalized, no limitations on activities (as recommended by the WHO R&D Blueprint Group).^26^ Statistical significance for differences in temperature, serum CRP concentration, serum IL-6 concentration, absolute lymphocyte counts (ALC), and platelet counts on day −1 versus day 3 post-lenzilumab was determined using a two-tailed paired t-test. Day 3 was determined as the last value for statistical analysis as data post day 3 were not available for more than 50% of this cohort.

## RESULTS

### Patients and baseline characteristics

Twelve patients received full treatment with 3 doses of lenzilumab administered 8 hours apart. The baseline demographic and clinical characteristics of these patients are summarized in Table 1. Eight patients (67%) were male; the median age was 65.0 years (range 29-81). Median BMI was 29 (range 22-42). Nine patients were white, 2 were Asian, and 1 American Indian/Native American. All patients had at least one comorbidity associated with poor outcomes. Seven (58%) had diabetes mellitus, 7 (58%) had hypertension, 6 (50%) had obesity (BMI > 30), 2 (17%) had chronic kidney disease, 2 (17%) had coronary artery disease and 1 (8%) was on immunosuppressive therapy with a history of kidney transplantation. Seven (58%) had underlying lung disease: 4 (33%) with obstructive sleep apnea, 2 (17%) with chronic obstructive pulmonary disease, and 1 (8%) with reactive airway disease.

**Table 1.** Demographics and baseline characteristics.

| **Table 1. Demographics and baseline characteristics** |  |
| --- | --- |
| **Characteristic** | **Patients (n=12)** |
| **Age, years** | 65 (52-70)* |
| **Gender** |  |
| Male | 8 (67%) |
| Female | 4 (33%) |
| **BMI** | 29 (24-36)* |
| **Race** |  |
| White | 9 (75%) |
| Asian | 2 (17%) |
| American Indian/Native Indian | 1 (8%) |
| **Comorbidities** |  |
| Diabetes mellitus | 7 (58%) |
| Hypertension | 7 (58%) |
| Obesity | 6 (50%) |
| Chronic kidney disease | 2 (17%) |
| Coronary artery disease | 2 (17%) |
| Kidney transplantation | 1 (8%) |
| Obstructive lung disease | 4 (33%) |
| Chronic obstructive pulmonary disease | 2 (17%) |
| Reactive airway disease | 1 (8%) |
| **Temperature, °C** | 38 (37.25-38.5)* |
| **Inflammatory markers before lenzilumab** |  |
| CRP ($\leq 8.0$ mg/L) | 103.2 (52.7-159.9)* |
| Ferritin (Males: 24-336 mcg/L,<br>Females: 11-307 mcg/L) | 596 (358.3-709.0)* |
| IL-6 ($\leq 1.8$ pg/mL) | 30.95 (24.18-34.05)* |
| D-dimer ($\leq 500$ ng/mL) | 829 (513.5-1298.5)* |
| **Lymphocyte count before lenzilumab (0.95 - 3.07 x 10<sup>9</sup>/L)** | 0.75 (0.55-1.04)* |
| **Oxygen therapy before lenzilumab** |  |
| High-flow oxygen | 3 (25%) |
| Nasal cannula | 8 (67%) |
| Invasive ventilation | 0 (0%) |
| Noninvasive ventilation | 1 (8%) |
| **SpO<sub>2</sub>/FiO<sub>2</sub> ratio before lenzilumab** | 280.9 (252.5-317.9)* |

\*Median (IQR)

All patients required oxygen supplementation at baseline; 1 patient was on non-invasive positive-pressure ventilation, 8 (67%) were on low flow oxygen, 3 (25%) were on high flow oxygen. The median SpO2/FiO2 ratio was 281, with SpO2/FiO2 ratios below 315 in 8 (67%) patients, and below 235 in 3 (25%) patients. Additionally, 6 (50%) patients were febrile within 24-48 hours prior to lenzilumab administration, with a median temperature of 38.3 °C.

Seven (58%) patients had lymphopenia at baseline, with an absolute lymphocyte count less than 0.95 × 10^9^/L. All patients had an elevation in at least one inflammatory marker at baseline. Eleven (92%) patients had elevated CRP values above the upper limit of normal (>8.0 mg/L), with a median of 103.2 mg/L. Ten (83%) patients had elevated ferritin values above the upper limit of normal (>336 mcg/L), with a median of 596 mcg/L. All 11 patients with IL-6 levels available at baseline had elevated values above the upper limit of normal (>1.8 pg/mL), with a median of 30.95 pg/mL. Of the 11 patients with D-dimer levels available at baseline, 9 (75%) had values above the upper limit of normal (>500 ng/mL), with a median of 829 ng/mL.

### Clinical Outcomes

Clinical improvement, as defined by the improvement of at least 2 points on the 8-point ordinal clinical endpoints scale, was observed in 11 out of 12 (92%) patients (Figure 1A), with a ≥ 3-point improvement in 10 patients and a 2-point improvement in 1 patient (Figure 1A). The median time to a 2-point clinical improvement was 5 days (95% CI, 2-7 days). All 11 patients with clinical improvement were discharged after a median of 5 days (range 3 −19) post-lenzilumab. The patient discharged on day 19 was ready for discharge on day 9 but remained hospitalized for social reasons.

**Figure 1.**
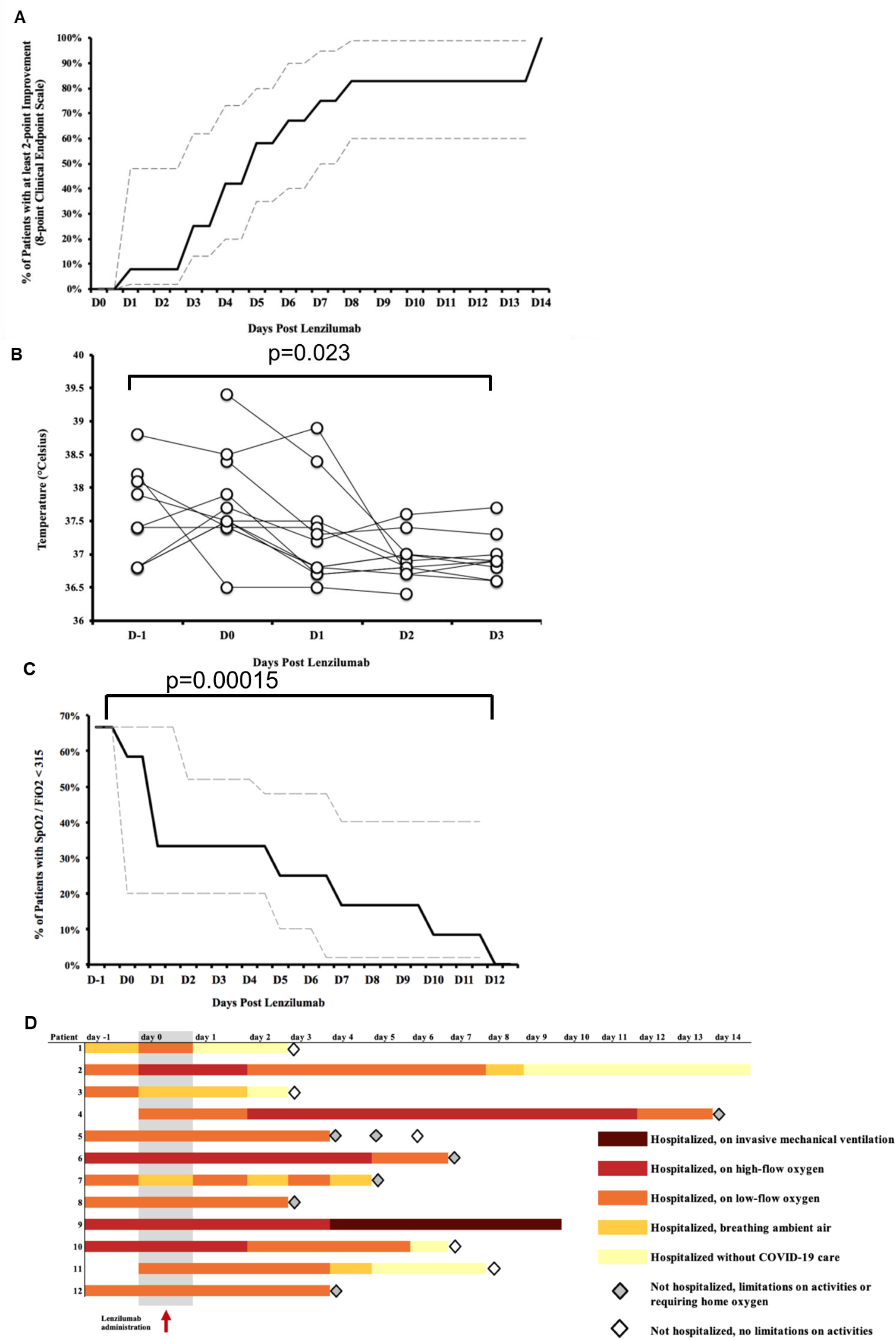
Lenzilumab treatment results in improved clinical outcomes of patients with severe and critical COVID-19 pneumonia. A) Cumulative percentage of patients with at least 2 point improvement in 8 point clinical endpoint scale (95% Kaplan Meier confidence interval displayed). B) Individual temperature over time post-lenzilumab treatment C) Percentage of patients with SpO2/FiO2 <315 over time post-lenzilumab treatment (95% Kaplan Meier confidence interval displayed) D) Individual hospitalization and oxygen requirement status.

There was a significant improvement in mean temperature at day 3 compared to baseline (37.95 vs. 36.97, p=0.023, Figure 1B). In patients who were febrile at baseline, fever resolved within 48 hours of lenzilumab administration. There was a significant improvement in the proportion of patients with SpO2/FiO2 < 315 at the end of observation compared to baseline (8% vs. 67%, p=0.00015 SpO2/FiO2 level baseline vs. last value, Figure 1C). Of 8 patients with SpO2/FiO2 < 315 at baseline, SpO2/FiO2 improved to > 315 in four on day 1 post-lenzilumab. Five (42%) patients were discharged on home oxygen, including one patient who had been on home oxygen pre-COVID-19 illness. One patient (8.3%) required invasive mechanical ventilation. There were no deaths. Figure 1D depicts individual patient hospitalization and oxygen requirement status.

### Laboratory markers

Compared to baseline, there was significant improvement in mean CRP and IL-6 on day 3 following lenzilumab administration (137.3 mg/L vs. 51.2 mg/L, p = 0.040; 26.8 pg/mL vs. 16.1 pg/mL, p = 0.035; respectively) (Figures 2A, B). Compared to baseline, an improvement of at least 50% was observed in CRP levels in 6 patients (50%) by day 2, and IL-6 levels in 4 patients (33.3%) by day 3. There was a significant increase in mean platelet count from baseline to day 3 post lenzilumab (217.7 ×10^9^/L vs 261.8 ×10^9^/L, p = 0.001, Figure 2C). There was also a trend toward improved absolute lymphocyte counts on day 3 compared to baseline (0.89 × 10^9^/L vs 1.14 × 10^9^/L, p = 0.107, Figure 2C). Analysis of human cytokines comparing pretreatment with 48 hours post-lenzilumab treatment in one patient revealed significant reduction in multiple cytokines involved in the cytokine storm (G-CSF, MDC, GM-CSF, IL-1α, IFN-γ, IL-7, FLT-3L, IL-1rα, IL-6, IL-12p70, Figure 2E).

**Figure 2.**
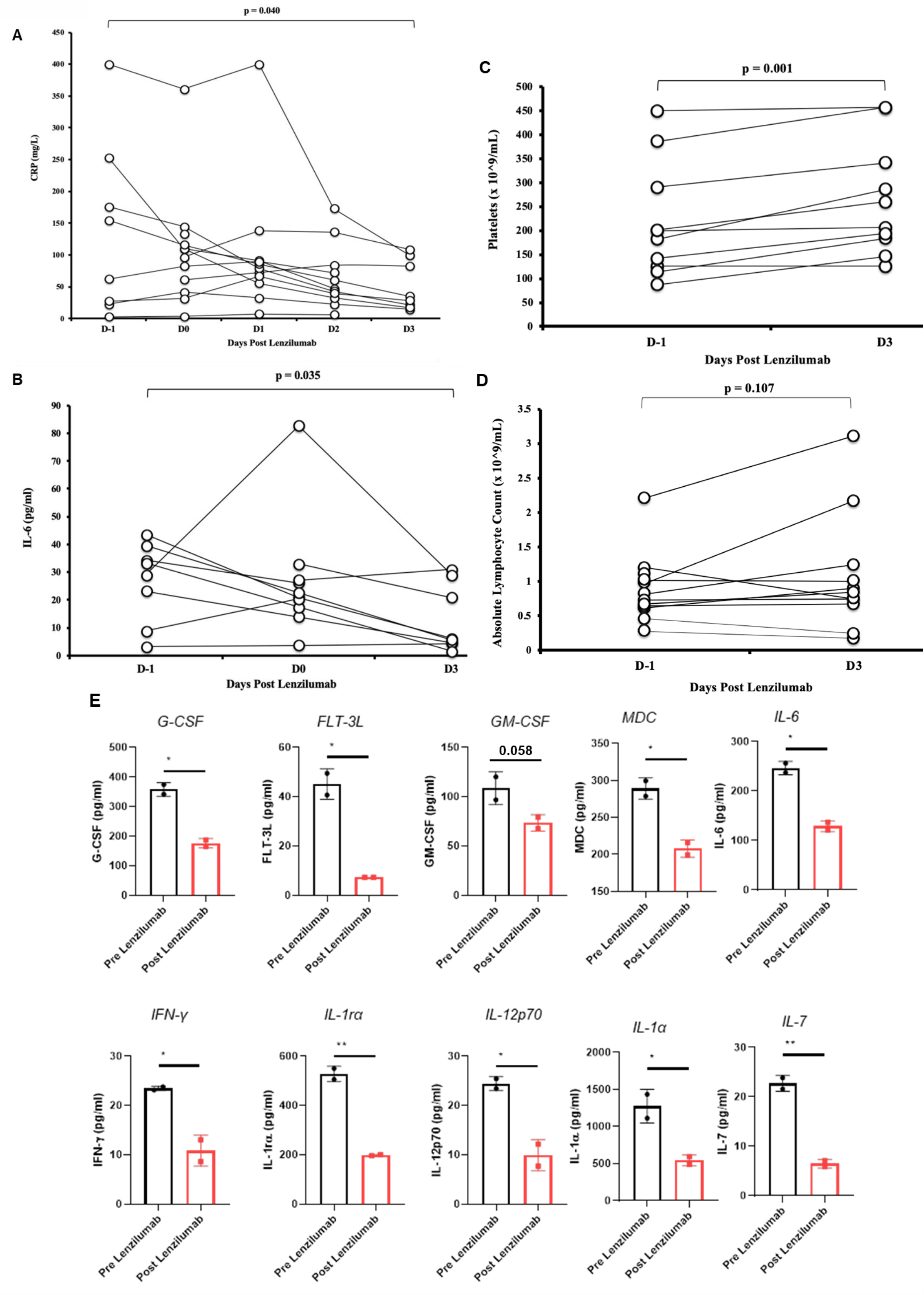
Lenzilumab treatment results in improved inflammatory cytokines and markers of disease severity in patients with severe and critical COVID-19 pneumonia. A) Individual CRP level over time post-lenzilumab treatment. B) Individual IL-6 levels, on Day −1, Day 0 and Day 3 post-lenzilumab treatment. C) Individual platelet levels on Day −1 and Day 3 post-lenzilumab treatment D) Individual absolute lymphocyte count on Day −1 and Day 3 post-lenzilumab treatment E) Inflammatory cytokine levels on Day −1 and Day 2 post-lenzilumab treatment.

### Safety of Lenzilumab Treatment

There was no significant difference in mean absolute neutrophil count or hemoglobin values between baseline and day 3 post lenzilumab: 5.1×10^9^/L vs. 4.8 ×10^9^/L, p=0.27; 12.9 g/dL vs. 11.4 g/dL, p=0.89; respectively. In one patient, hemoglobin values dropped from 10.3 g/dL on day 0 to 7.9 g/dL on day 6. This patient had undergone a renal biopsy on day 2; imaging revealed a subcapsular hematoma. At the last study observation, the patient remained anemic at 9.3 g/dL. There were no infusion reactions with lenzilumab administration. One patient, with a history of restless leg syndrome, reported a “pins and needles” sensation during the first dose of lenzilumab; those symptoms resolved and did not recur with subsequent infusions of lenzilumab. No other treatment-emergent adverse events attributable to lenzilumab were noted.

## DISCUSSION

There is no therapy with proven efficacy against COVID-19 at present. We report our observations from the first-ever use of lenzilumab to neutralize GM-CSF in the treatment of COVID-19. Lenzilumab was offered through a compassionate single-use IND to patients with severe and critical COVID-19 pneumonia. Based on the pathophysiology of cytokine storm following SARS-CoV-2 infection^1,27^, along with our preclinical work^21^, we hypothesized that lenzilumab-induced GM-CSF depletion prevents CRS in COVID-19 and progression to severe disease or death. At baseline, all 12 patients had at least one risk factor associated with poor outcomes: age, smoking history, cardiovascular disease, diabetes, chronic kidney disease, chronic lung disease, high BMI, and elevated inflammatory markers, with several patients having multiple such risk factors.^28,29^ In this cohort of high-risk patients with severe and critical COVID-19 pneumonia, treatment with lenzilumab was associated with improved overall clinical outcome in 11/12 patients (91.7%) on an 8-point ordinal scale; all 11 patients were discharged after a median of 5 days. Significant improvement in oxygen requirement, as well as inflammatory cytokines and markers of disease severity, were also observed. These results are consistent with our original hypothesis, and corroborate our laboratory findings following GM-CSF depletion in preclinical models of CRS after CART cell therapy.^21^ In addition, the use of lenzilumab was associated with a significant improvement in platelet count, indicating possibly an overall improved coagulopathy associated with CRS post-COVID-19.^30^ Interestingly, the use of lenzilumab in this cohort was associated with a trend to an increase in lymphocyte count (Figure 2D). We have recently shown that GM-CSF depletion results in modulation of apoptosis pathways in T cells.^22^ It is unclear at this time if the increase in lymphocyte count is secondary to clearance of SARS-CoV-2 virus, or a direct effect of GM-CSF on T cells; this question will be answered in the planned phase III trial. Figure 3 depicts a proposed mechanism for the role of GM-CSF in CRS post-COVID-19.

**Figure 3.**
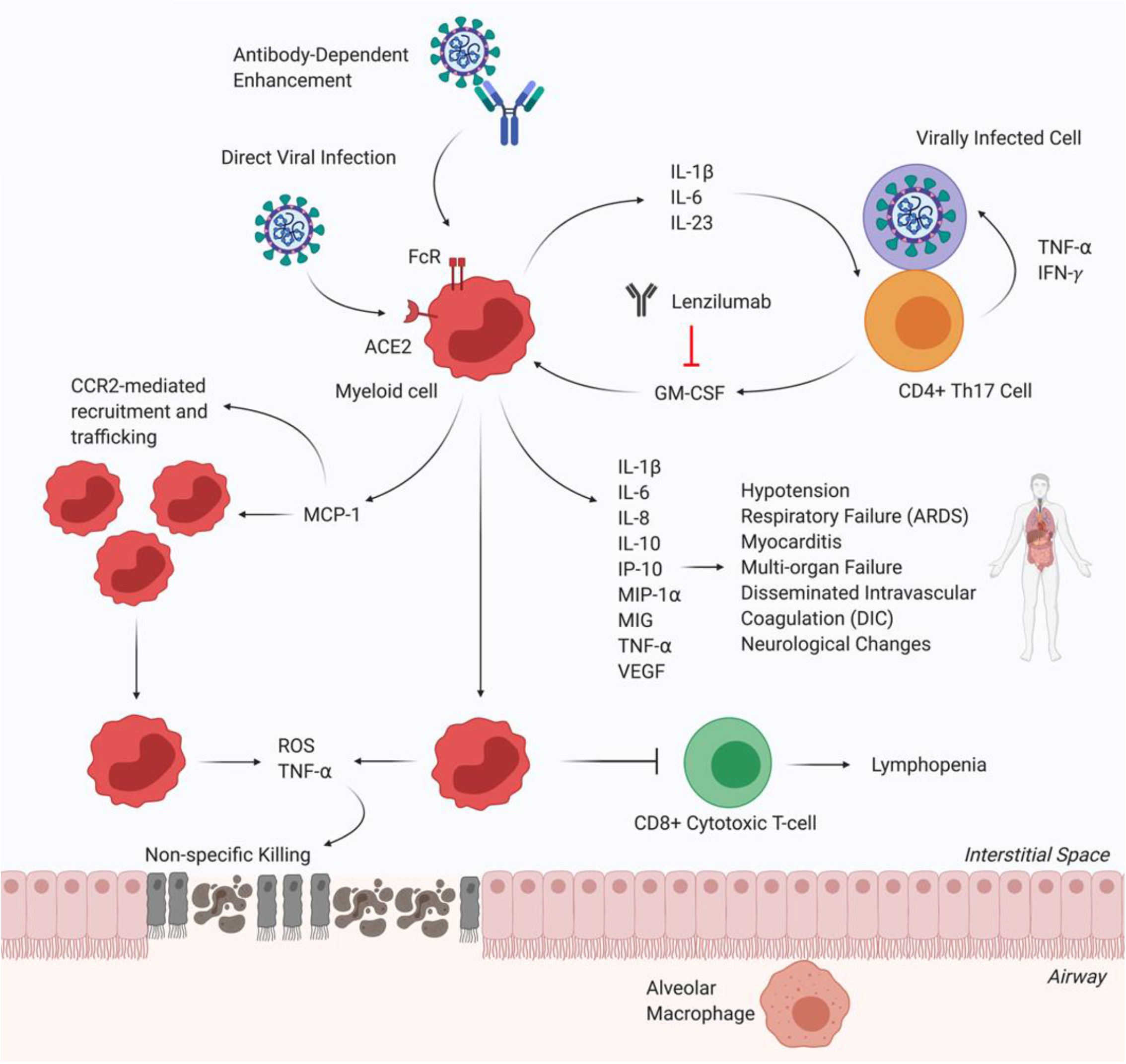
Proposed mechanim for GM-CSF depletion in COVID-19 associated cytokine storm.

Five patients received other pharmacotherapies targeting COVID-19 besides lenzilumab. Three patients received hydroxychloroquine; one patient received remdesivir and one patient received steroids. Two patients received lenzilumab after the failure of clinical improvement with either hydroxychloroquine or remdesivir and subsequently improved. Two patients received lenzilumab concomitantly with hydroxychloroquine; both of these patients were discharged home. One of these patients also received off-label tocilizumab on day 6 post-lenzilumab and was released on home oxygen. One patient received steroid therapy concomitantly with lenzilumab; this patient remained on invasive mechanical ventilation on the last day of observation.

The use of lenzilumab was safe, without any adverse events attributable to lenzilumab. While there is a theoretical concern for bone marrow toxicity when GM-CSF is depleted, lenzilumab treatment was not associated with any hematological toxicity in this cohort. There were no infusion reactions following lenzilumab treatment. Importantly, a sensation of pins and needles reported by one patient while receiving lenzilumab, did not recur with subsequent infusions; the patient had a history of restless leg syndrome. Restless legs have not been described in any of the non-COVID-19 patients who have received lenzilumab for other indications.

Our report has several limitations. First, the sample size is small and did not include controls. Second, as lenzilumab was offered under emergency single-use IND conditions, all management decisions, including prescribing medications and laboratory/radiologic monitoring, were at the discretion of the treating clinicians. This resulted in some heterogeneity in the treatment specifics of individual patients as well as the laboratory and other diagnostic data that were collected. Given this and the absence of a control arm in the study, we cannot, with full confidence, declare that the clinical improvement that we noted in our patients was clearly and solely attributable to lenzilumab. These limitations will be addressed in the recently initiated randomized Phase III clinical trial (NCT04314843).

In summary, we administered lenzilumab, under a single-use emergency IND compassionate program, to 12 patients with severe and critical COVID-19 pneumonia and with risk factors for disease progression. Lenzilumab use was associated with improved clinical outcomes, oxygen requirement, and cytokine storm in this cohort of patients, with no reported mortality. Lenzilumab was well tolerated; no treatment-emergent adverse events attributable to lenzilumab were observed.

## Data Availability

All data is included in the manuscript

## Acknowledgements

Lenzilumab was provided by Humanigen, Inc. This work was supported through grants from K12CA090628 (SSK), Mayo Clinic K2R Career Development Program (SSK), ADB is supported by Grants AI 110173 and AI120698 from NIAID, 109593-62-RGRL from Amfar, and the HH Sheikh Khalifa Bin Zayed Al-Nahyan named professorship from Mayo Clinic.

## REFERENCES

1. Huang C, Wang Y, Li X, et al. Clinical features of patients infected with 2019 novel coronavirus in Wuhan, China. Lancet 2020;395:497–506.

2. Chen N, Zhou M, Dong X, et al. Epidemiological and clinical characteristics of 99 cases of 2019 novel coronavirus pneumonia in Wuhan, China: a descriptive study. Lancet 2020;395:507–13.

3. Moore JB, June CH. Cytokine release syndrome in severe COVID-19. Science 2020;368:473–4.

4. Teachey DT, Lacey SF, Shaw PA, et al. Identification of Predictive Biomarkers for Cytokine Release Syndrome after Chimeric Antigen Receptor T-cell Therapy for Acute Lymphoblastic Leukemia. Cancer discovery 2016;6:664–79.

5. June CH, Sadelain M. Chimeric Antigen Receptor Therapy. N Engl J Med 2018;379:64–73.

6. Schram AM, Berliner N. How I treat hemophagocytic lymphohistiocytosis in the adult patient. Blood 2015;125:2908–14.

7. Neelapu SS, Locke FL, Bartlett NL, et al. Axicabtagene Ciloleucel CAR T-Cell Therapy in Refractory Large B-Cell Lymphoma. N Engl J Med 2017;377:2531–44.

8. Maude SL, Laetsch TW, Buechner J, et al. Tisagenlecleucel in Children and Young Adults with B-Cell Lymphoblastic Leukemia. N Engl J Med 2018;378:439–48.

9. Brudno JN, Kochenderfer JN. Toxicities of chimeric antigen receptor T cells: recognition and management. Blood 2016.

10. Hay KA, Turtle CJ. Chimeric Antigen Receptor (CAR) T Cells: Lessons Learned from Targeting of CD19 in B-Cell Malignancies. Drugs 2017.

11. Fitzgerald JC, Weiss SL, Maude SL, et al. Cytokine Release Syndrome After Chimeric Antigen Receptor T Cell Therapy for Acute Lymphoblastic Leukemia. Critical Care Medicine 2017;45:e124–e31.

12. Gust J, Hay KA, Hanafi LA, et al. Endothelial Activation and Blood-Brain Barrier Disruption in Neurotoxicity after Adoptive Immunotherapy with CD19 CAR-T Cells. Cancer Discovery 2017;7:1404–19.

13. Li X, Wang L, Yan S, et al. Clinical characteristics of 25 death cases with COVID-19: A retrospective review of medical records in a single medical center, Wuhan, China. Int J Infect Dis 2020;94:128–32.

14. Wang F, Hou H, Luo Y, et al. The laboratory tests and host immunity of COVID-19 patients with different severity of illness. JCI Insight 2020.

15. Arnaldez FI, O’Day SJ, Drake CG, et al. The Society for Immunotherapy of Cancer perspective on regulation of interleukin-6 signaling in COVID-19-related systemic inflammatory response. J Immunother Cancer 2020;8.

16. Ascierto PA, Fox B, Urba W, et al. Insights from immuno-oncology: the Society for Immunotherapy of Cancer Statement on access to IL-6-targeting therapies for COVID-19. J Immunother Cancer 2020;8.

17. Henry BM, de Oliveira MHS, Benoit S, Plebani M, Lippi G. Hematologic, biochemical and immune biomarker abnormalities associated with severe illness and mortality in coronavirus disease 2019 (COVID-19): a meta-analysis. Clin Chem Lab Med 2020.

18. Zhou Y, Fu B, Zheng X, et al. Aberrant pathogenic GM-CSF<sup>+</sup> T cells and inflammatory CD14<sup>+</sup>CD16<sup>+</sup> monocytes in severe pulmonary syndrome patients of a new coronavirus. bioRxiv 2020:2020.02.12.945576.

19. Xiong Y, Liu Y, Cao L, et al. Transcriptomic characteristics of bronchoalveolar lavage fluid and peripheral blood mononuclear cells in COVID-19 patients. Emerg Microbes Infect 2020;9:761–70.

20. Xu X, Han M, Li T, et al. Effective treatment of severe COVID-19 patients with tocilizumab. Proc Natl Acad Sci U S A 2020.

21. Sterner RM, Sakemura R, Cox MJ, et al. GM-CSF inhibition reduces cytokine release syndrome and neuroinflammation but enhances CAR-T cell function in xenografts. Blood 2019;133:697–709.

22. Cox MJ, Kuhlmann CJ, Sterner RM, et al. Improved anti-tumor response of chimeric antigen receptor t cell (CART) therapy after GM-CSF inhibition is mechanistically supported by a novel direct interaction of GM-CSF with activated CARTs. Blood 2019;2019:3868.

23. Padron E, Painter JS, Kunigal S, et al. GM-CSF-dependent pSTAT5 sensitivity is a feature with therapeutic potential in chronic myelomonocytic leukemia. Blood 2013;121:5068–77.

24. Patnaik MM, Sallman DA, Mangaonkar A, et al. Phase 1 study of lenzilumab, a recombinant anti-human GM-CSF antibody, for chronic myelomonocytic leukemia (CMML). Blood 2020.

25. Molfino NA, Kuna P, Leff JA, et al. Phase 2, randomised placebo-controlled trial to evaluate the efficacy and safety of an anti-GM-CSF antibody (KB003) in patients with inadequately controlled asthma. BMJ Open 2016;6:e007709.

26. WHO R&D Blueprint: novel Coronavirus, COVID-19 Therapeutic Trial Synopsis. https://www.who.int/blueprint/priority-diseases/key-action/COVID-19_Treatment_Trial_Design_Master_Protocol_synopsis_Final_18022020.pdf

27. Merad M, Martin JC. Pathological inflammation in patients with COVID-19: a key role for monocytes and macrophages. Nature Reviews Immunology 2020.

28. Jordan RE, Adab P, Cheng KK. Covid-19: risk factors for severe disease and death. BMJ 2020;368:m1198.

29. Wu Z, McGoogan JM. Characteristics of and Important Lessons From the Coronavirus Disease 2019 (COVID-19) Outbreak in China: Summary of a Report of 72314 Cases From the Chinese Center for Disease Control and Prevention. JAMA 2020.

30. McGonagle D, O’Donnell JS, Sharif K, Emery P, Bridgewood C. Immune mechanisms of pulmonary intravascular coagulopathy in COVID-19 pneumonia. Lancet Rheumatology 2020.

